# Brain white matter after pediatric mild traumatic brain injury: a diffusion tensor and neurite orientation and dispersion imaging study

**DOI:** 10.1101/2021.09.09.21263356

**Authors:** Ayushi Shukla, Ashley L. Ware, Sunny Guo, Bradley Goodyear, Miriam H. Beauchamp, Roger Zemek, William Craig, Quynh Doan, Christian Beaulieu, Keith O. Yeates, Catherine Lebel, on behalf of the Pediatric Emergency Research Canada A-CAP study team

## Abstract

**Background:** Pediatric mild traumatic brain injury (mTBI) affects millions of children annually. Diffusion tensor imaging (DTI) is sensitive to axonal injuries and white matter microstructure and has been used to characterize the brain changes associated with mild traumatic brain injury (mTBI). Neurite orientation dispersion and density imaging (NODDI) is a diffusion model that can provide additional insight beyond traditional DTI metrics, but has not been examined in pediatric mTBI. The goal of this study was to employ DTI and NODDI to gain added insight into white matter alterations in children with mTBI compared to children with mild orthopedic injury (OI).

**Methods:** Children (mTBI n=320, OI n=176) aged 8-16.99 years (*m* 12.39 ± 2.32 years) were recruited from emergency departments at five hospitals across Canada and underwent 3T MRI on average 11 days post-injury. DTI and NODDI metrics were calculated for seven major white matter tracts and compared between groups using univariate analysis of covariance controlling for age, sex, and scanner type. False discovery rate (FDR) was used to correct for multiple comparisons.

**Results:** Univariate analysis revealed no significant group main effects or interactions in DTI or NODDI metrics. Fractional anisotropy and neurite density index in all tracts exhibited a significant positive association with age and mean diffusivity in all tracts exhibited a significant negative association with age in the whole sample.

**Conclusions:** Overall, there were no differences between mTBI and OI groups in brain white matter microstructure from either DTI or NODDI in the seven tracts. This indicates that mTBI is associated with relatively minor white matter differences, if any, at the post-acute stage. Brain differences may evolve at later stages of injury, so longitudinal studies with long-term follow-up are needed.

## 1. Introduction

Traumatic brain injury (TBI) is a global public health concern that affects millions of children annually.^1^ Seventy-five to ninety percent of all TBIs are mild in severity.^2,3^ Mild TBI can be associated with ongoing emotional, cognitive, and physical complaints, such as headaches, irritability, and forgetfulness, known as persistent post-concussive symptoms (PPCS).^4–6^ In a majority of children with mild TBI, symptoms resolve within 4-6 weeks of injury.^7,8^ Yet, up to 30% of injured children continue to experience persistent post concussive symptoms (PPCS) one month or more after injury.^9,10^

The rotational and shearing forces from mTBIs can lead to axonal injuries,^11,12^ which may underlie PPCS and negative outcomes after mTBI.^13^ These injuries are usually not detectible using conventional imaging techniques such as computed tomography,^14^ but techniques, such as diffusion MRI, more specifically, diffusion tensor imaging (DTI), offer promise for greater sensitivity.^12^ Generally, post-acutely (4-20 days post injury), higher fractional anisotropy (FA) and lower mean diffusivity (MD) are observed in children with mTBI as compared to healthy controls, indicating possible edema,^15–21^ but findings have been mixed, with some studies finding no changes^22^ or lower FA.^23,24^ Altered FA, globally as well as in specific tracts, at 3-12 months post-injury have been identified in symptomatic children with mTBI, but not asymptomatic children.^19,26^

Neurite orientation dispersion and density imaging (NODDI)^27^ is another diffusion model that provides three metrics with potentially more specificity than traditional DTI: 1) neurite density index (NDI), sensitive to myelin and axonal density,^28^ 2) orientation dispersion index (ODI), a measure of angular variation or fanning of neurites,^29^ and 3) fraction of the isotropic diffusion compartment (FISO), an estimate of the free water content in the brain that corresponds to the cerebrospinal fluid space.^27^ No published studies have examined NODDI metrics in pediatric mTBI, but studies in adults have yielded promising results. Adults with mTBI showed lower NDI and higher FISO as compared to those with orthopedic injuries, predominantly in anterior brain regions, at two weeks post injury, with subsequent decreases in NDI longitudinally^30^. In a longitudinal follow-up of concussed college athletes, spatially extensive decreases in NDI and an increase in ODI over time were identified.^31^ However, pediatric mTBI presents unique physiological characteristics and different recovery mechanisms compared to adults.^19,32,33^ Children have distinct cerebral metabolisms, different cranial anatomies and biomechanical properties of injury. In addition, neurobehavioral outcomes after mTBI are a confluence of environmental, age, and psychiatric factors.^34–37^ These differences highlight the need for neuroimaging studies of mTBI using NODDI specifically in pediatric samples.

The current study sought to investigate white matter alterations following mTBI in children 8-16 years of age at the post-acute stage (2-33 days) of injury using DTI and NODDI as compared to an orthopedic injury (OI) comparison group. We hypothesized that children with mTBI would exhibit higher FA and lower MD in several white matter tracts compared to the OI comparison group. We also expected lower NDI and higher FISO post-injury in the mTBI group as compared to the OI group.

## 2. Methods

### 2.1. Study design and procedure

Data were collected as part of Advancing Concussion Assessment in Pediatrics (A-CAP), a larger multi-site longitudinal cohort study of mTBI.^38^ Children aged 8 to 16.99 years (mean 12.39 ± 2.32) were recruited within 48 hours of mTBI or OI in the emergency department (ED) of five hospitals that are members of the Pediatric Emergency Research Canada (PERC) network^39^: Alberta Children’s Hospital (Calgary), Children’s Hospital of Eastern Ontario (Ottawa), Centre Hospitalier Universitaire (CHU) Sainte-Justine (Montreal), Stollery Children’s Hospital (Edmonton), and British Columbia Children’s Hospital (Vancouver). Children returned for a post-acute assessment that included diffusion MRI around 10 (mean 11.56 ± 5.43) days post-injury. The study was approved by the research ethics board at each site and informed consent/assent was obtained from participants and their parents and guardians. The detailed protocol for the study has been published elsewhere^38^.

### 2.2. Participants

To be eligible for the study, children had to present to the ED within 48 hours of injury and at least one parent and the child had to speak and understand English (or French in Montreal and Ottawa). A total of 967 participants were recruited from the ED, 827 returned for the post-acute follow-up, and 628 completed MRI, and scans for 496 children (mTBI n=320, OI n=176) passed quality assurance procedures which included examining the MRIs for motion related and other artefacts and were included in the current study. The participants who completed post-acute MRI did not differ in terms of age, sex, or socio-economic status (SES) compared to children who did not return for post-acute follow-up or who returned but did not complete MRI.

#### 2.2.1. Mild TBI

The mTBI group included children who had a blunt head trauma resulting in one or more of the following three criteria: 1) observed loss of consciousness, 2) a Glasgow coma scale^40^ score of 13-15, or 3) one or more acute signs/symptoms of concussion (i.e., post-traumatic amnesia, post traumatic seizure, vomiting, headaches, dizziness, changes in mental status) identified in the ED by the medical personnel on a case report form.

#### 2.2.2. Mild OI

The OI group met the following inclusion criteria: 1) fracture, sprain, or strain to the upper or lower extremity from a blunt force trauma, and 2) an Abbreviated Injury Scale^41^ (AIS) score of 4 or less. An OI comparison group helps control for premorbid demographic and behavioral risk factors (e.g., attention deficit/hyperactivity disorder (ADHD) and impulsivity are associated with higher risk of injury^42,43^), as well as for sequelae of injury that are not specific to mild TBI (such as pain, requirement of rest), and thus may better delineate the specific effects of mild TBI on brain white matter structure than a healthy comparison group.

#### 2.2.3. Exclusion criteria

Exclusion criteria for the mTBI group were as follows: 1) delayed neurological deterioration (GCS <13), 2) need for neurosurgical intervention, or 3) loss of consciousness for more than 30 minutes or post-traumatic amnesia >24 hours. For the mild OI group, exclusion criteria were: 1) Any evidence of head trauma or concussion; 2) injuries requiring surgical intervention or procedural sedation. Both groups had the following exclusion criteria: 1) hypoxia hypotension, or shock during or following the injury, 2) previous TBI requiring overnight hospital stay, 3) previous concussion within 3 months, 4) pre-existing neurological or neurodevelopmental disorders, 5) hospitalisation for psychiatric deficits within the previous 1 year, 6) sedative medication administered during ED visit, 7) injury accompanied by alcohol and/or drug use, 8) abuse or assault related injuries, 9) absence of legal guardians or child in foster care. Children with contraindications to MRI were included in A-CAP but excluded from the present study because they could not complete MRI.

### 2.3. Magnetic Resonance Imaging

Participants completed T1-weighted and diffusion imaging at the post-acute assessment between 2-23 days (mean 11.56 ± 5.43) post injury. Images were acquired on a 3T scanner at each site (General Electric MR750w in Calgary; General Electric MR750 and Siemens Prisma in Montreal, General Electric MR750 in Vancouver; Siemens Prisma in Edmonton; Siemens Skyra in Ottawa).

#### 2.3.1. T1 Acquisition

3D T1-weighted magnetisation prepared rapid acquisition gradient echo (MP RAGE)/ Fast spoiled gradient echo brain volume (FSPGR BRAVO) images with repetition time (TR) = 1880/8.25 ms, echo time (TE) = 2.9/3.16 ms, inversion time (TI) = 948/600 ms, field of view (FOV) = 25.6/24 cm, resolution = 0.8x0.8x0.8 mm isotropic, number of slices = 192, and a flip angle of 10° were acquired for sites with Siemens/GE scanners respectively.

#### 2.3.2. DTI Acquisition

Spin-echo, single-shot echo planar imaging (EPI) was used to acquire DTI images with 5 b = 0 s/mm^2^, 30 gradient directions at b = 900 s/mm^2^, 30 gradient directions at b = 2000 s/mm^2^, FOV = 22/24.2 cm, TR = 6300/12000 ms, TE = 55/98 ms, and a resolution of 2.2 mm isotropic at sites with Siemens/GE scanners respectively.

#### 2.3.3. Quality Control

The T1-image data were manually rated for motion by two trained analysts using a 0-2 ordinal scale with “0” assigned to images with gross artifacts that were considered *unusable*, a rating of “1” assigned to images with apparent, but minor artifacts that were *acceptable* for use, and a “2” assigned to images that were free from visible artifact and were considered to be of *excellent* quality.

For diffusion weighted images, scans with more than 7 (i.e., > 25%) volumes rated unusable, for b900 and b2000 image each, were discarded. For scans with fewer than 8 volumes with motion artifacts, the volumes with motion were removed before calculation of diffusion parameters, but the participants were still included in analysis. For an image to be considered usable, it needed to have atleast 1 or more usable b0.

#### 2.3.4. Image Processing

T1- and diffusion-weighted DICOM data were converted into NIfTI format using the dcm2niix tool in MRIcron (publicly available software; https://github.com/rordenlab/dcm2niix), and the bval and bvec files were automatically created from the raw diffusion-weighted DICOM headers. During conversion to NIfTI format, T_1_-weighted images were automatically reoriented to canonical space.

T1-weighted images were processed on the Advanced Remote Cluster (ARC), a remote Linux computing cluster at the University of Calgary, AB, Canada. Brain extractions of T1-weighted images were obtained using the Advanced Normalization Tools version 3.0.0.0.dev13-ga16cc (compiled January 18, 2019) volume-based cortical thickness estimation pipeline (antsCorticalThickness.sh), with the OASIS pediatric template from the MICCAI 2012 Multi Atlas Challenge ^44^ used for anatomical reference during skull-stripping.

The diffusion-weighted images (b900 and b2000) were corrected for eddy current distortions, motion artifact, and Gibbs ringing, and were tensor fitted using ExploreDTI v4.8.6 running on MATLAB v8.6.0 R2018a (MathWorks Inc., Natick, MA). Semi-automated deterministic streamline tractography^45^ was performed to delineate fiber tracts from the arcuate fasciculus (AF), cingulum bundle, cortico-spinal tract (CST), corpus callosum (CC), inferior fronto-occipital fasciculus (IFOF), inferior longitudinal fasciculus (ILF), and uncinate fasciculus (UF). DTI and NODDI metrics for each tract were averaged across the left and right hemisphere. The FA map of a representative participant was used to register FA maps of all other participants. All tract regions of interest were drawn on this representative participant and registered to other participants’ native space data, in which tractography was performed. Average fractional anisotropy (FA), Mean Diffusivity (MD) values for each tract were calculated for each participant.

Preprocessed data from the ExploreDTI toolbox were exported to the MATLAB NODDI Toolbox (http://www.nitrc.org/projects/noddi_toolbox) and fitted to the NODDI model, obtaining maps of intracellular (f_icvf_ or NDI) and isotropic (f_iso_) volume fractions and orientation diffusion index (ODI) for each brain tract for which DTI metrics were obtained.

#### 2.3.5. Statistical Analysis

Demographic data for the sample was examined using analyses of variance (ANOVA) and chi-square tests for continuous and categorical variables, respectively. Univariate analyses of covariance (ANCOVA) were performed to evaluate the relation between white matter metrics (FA, MD, NDI, ODI, FISO) and injury group (mTBI, OI), age, and sex, and their interactions. MRI scanner type was included in the models as a covariate. Non-significant interactions were trimmed from the final models. To correct for 7 multiple comparisons, false discovery rate (FDR)^46^ was used.

Power analysis using G*power 3.1^47^ revealed that at a small (*f* = .10), medium (*f* = .25), and large (*f* = .40) effect size *f* our study has a power of .66, .99, 1.00 respectively at critical *F*=3.85 with *α* = 0□05 with our sample size *n* = 560.

## 3. Results

Sample demographics are presented in *Table 1*. The groups did not differ in age, sex, or Full-Scale IQ. Mean and standard deviations of DTI (FA, MD) and NODDI (NDI, ODI, FISO) metrics for each group (mTBI and OI) are listed in *Table 2*. Metric maps for each metric for a representative study participant are depicted in *Figure 1*.

**Table 1.** Demographic data and injury characteristics for sample included in final analysis in the current study

| Variable |  | OI<br>*n* = 176 | TBI<br>*n* = 320 | p-value |
| --- | --- | --- | --- | --- |
| Age [Mean (SD)] |  | 12.5 (2.2)<br>(8.05-16.95) | 12.4 (2.4)<br>(8.00-16.96) | 0.619 |
| Full Scale IQ [Mean (SD)] * |  | 107 (13) | 106 (14) | 0.120 |
| Sex (%) | Female | 80 (45.5) | 120 (37.6) | 0.089 |
| PCS |  |  |  |  |
|  | Premorbid Somatic | 1.92 (2.79) | 3.05 (4.02) | .001 |
|  | Premorbid Cognitive | 7.05 (7.03) | 9.43 (7.74) | .001 |
|  | Post-acute Somatic | 1.38 (2.23) | 6.89 (5.46) | <.001 |
|  | Post-acute Cognitive | 6.21 (6.85) | 11.64 (.48) | <.001 |
| Race/Ethnicity (%) |  |  |  |  |
|  | White | 115 (65.3) | 214 (67.1) |  |
|  | Asian | 12 (6.8) | 27 (8.5) |  |
|  | Black | 4 (2.3) | 13 (4.1) |  |
|  | Hispanic | 8 (4.5) | 8 (2.5) |  |
|  | Indigenous | 3 (1.7) | 7 (2.2) |  |
|  | Other/Multi-Racial | 28 (15.9) | 43 (13.5) |  |

SD = Standard Deviation; \*collected only at 3-month follow-ups (*n* = 431); PCS = Post concussive symptoms

**Table 2.** Table depicting means and standard deviations of DTI (FA, MD) and NODDI (NDI, ODI, FISO) metrics for each tract for both groups (mTBI or OI) examined in this study.

| Brain Tract | DTI/NODDI metric | mTBI |  | OI |  |
| --- | --- | --- | --- | --- | --- |
|  |  | Mean | Standard deviation | Mean | Standard deviation |
| AF |  |  |  |  |  |
|  | FA | .44 | .04 | .45 | .04 |
|  | MD (mm<sup>2</sup>/s) | .78x10-3 | .25x10-4 | .77x10<sup>-3</sup> | .23x10<sup>-4</sup> |
|  | NDI | .55 | .05 | .55 | .05 |
|  | ODI | .26 | .04 | .27 | .03 |
|  | FISO | .07 | .03 | .07 | .04 |
| ILF |  |  |  |  |  |
|  | FA | .45 | .03 | .46 | .03 |
|  | MD (mm<sup>2</sup>/s) | .83x10<sup>-3</sup> | .27x10<sup>-4</sup> | .84x10<sup>-3</sup> | .25x10<sup>-4</sup> |
|  | NDI | .52 | .04 | .53 | .05 |
|  | ODI | .25 | .04 | .25 | .03 |
|  | FISO | .08 | .03 | .09 | .03 |
| IFOF |  |  |  |  |  |
|  | FA | .46 | .03 | .47 | .03 |
|  | MD (mm<sup>2</sup>/s) | .83x10<sup>-3</sup> | .25x10<sup>-4</sup> | .82x10<sup>-3</sup> | .24x10<sup>-4</sup> |
|  | NDI | .52 | .04 | .53 | .04 |
|  | ODI | .24 | .03 | .24 | .02 |
|  | FISO | .07 | .03 | .08 | .03 |
| Uncinate |  |  |  |  |  |
|  | FA | .41 | .03 | .41 | .03 |
|  | MD (mm<sup>2</sup>/s) | .84x10<sup>-3</sup> | .23x10<sup>-4</sup> | .83x10<sup>-3</sup> | .22x10<sup>-4</sup> |
|  | NDI | .46 | .04 | .47 | .05 |
|  | ODI | .26 | .03 | .26 | .03 |
|  | FISO | .05 | .04 | .05 | .05 |
| Cingulum |  |  |  |  |  |
|  | FA | .44 | .04 | .44 | .03 |
|  | MD (mm<sup>2</sup>/s) | .80x10<sup>-3</sup> | .28x10<sup>-4</sup> | .80x10<sup>-3</sup> | .28x10<sup>-4</sup> |
|  | NDI | .53 | .05 | .54 | .06 |
|  | ODI | .28 | .03 | .28 | .03 |
|  | FISO | .08 | .04 | .08 | .05 |
| CST |  |  |  |  |  |
|  | FA | .52 | .03 | .52 | .04 |
|  | MD (mm<sup>2</sup>/s) | .78x10<sup>-3</sup> | .37x10<sup>-4</sup> | .78x10<sup>-3</sup> | .38x10<sup>-4</sup> |
|  | NDI | .59 | .03 | .59 | .04 |
|  | ODI | .22 | .05 | .22 | .02 |
|  | FISO | .09 | .03 | .09 | .02 |
| CC |  |  |  |  |  |
|  | FA | .52 | .04 | .53 | .03 |
|  | MD (mm<sup>2</sup>/s) | .85x10<sup>-3</sup> | .36x10<sup>-4</sup> | .85x10<sup>-3</sup> | .35x10<sup>-4</sup> |
|  | NDI | .55 | .04 | .55 | .04 |
|  | ODI | .22 | .06 | .22 | .02 |
|  | FISO | .11 | .03 | .11 | .03 |

DTI = Diffusion tensor imaging, NODDI = Neurite orientation dispersion and density index, AF = Arcuate fasciculus, ILF = Inferior longitudinal fasciculus, IFOF = Inferior fronto-occipital fasciculus, CST = Corticospinal tract, CC = Corpus callosum, FA = Fractional anisotropy, MD = Mean diffusivity, NDI = Neurite density index, ODI = Orientation dispersion index, FISO = Fraction of isotropic water

**Figure 1:**
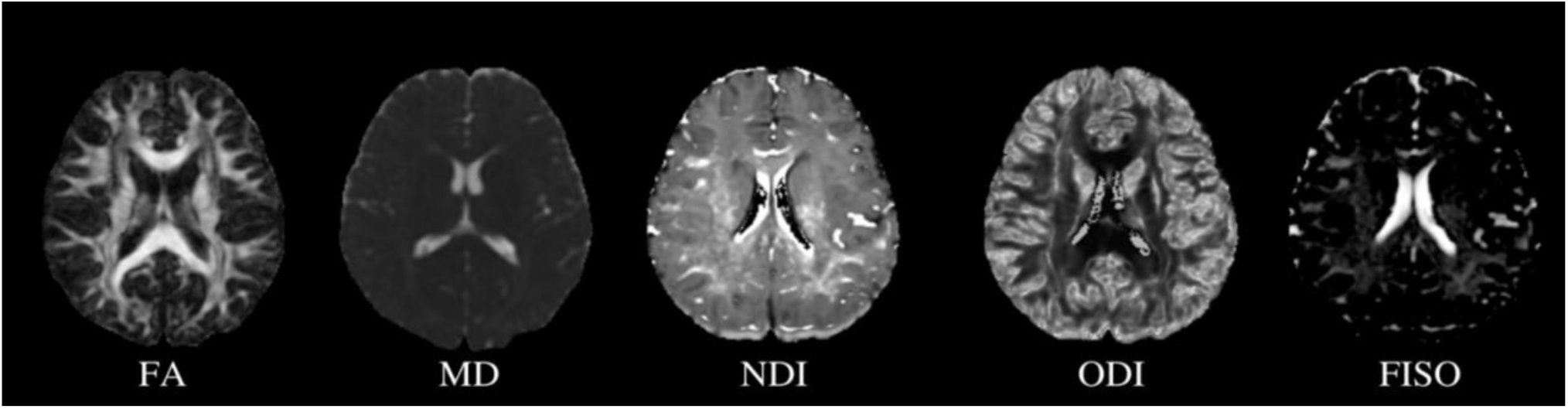
DTI (FA, MD) and NODDI (NDI, ODI, FISO) metric maps for a representative study participant. DTI = Diffusion tensor imaging, NODDI = Neurite orientation dispersion and density index, FA = Fractional anisotropy, MD = Mean diffusivity, NDI = Neurite density index, ODI = Orientation dispersion index, FISO = Fraction of isotropic water

### 3.1. DTI

Univariate ANCOVA results examining group differences in DTI metrics (FA, MD, RD, AD) are presented in *Table 3*. Children with mTBI or OI did not significantly differ on any DTI metric for any of the tracts examined in the current study after FDR correction.

**Table 3.**
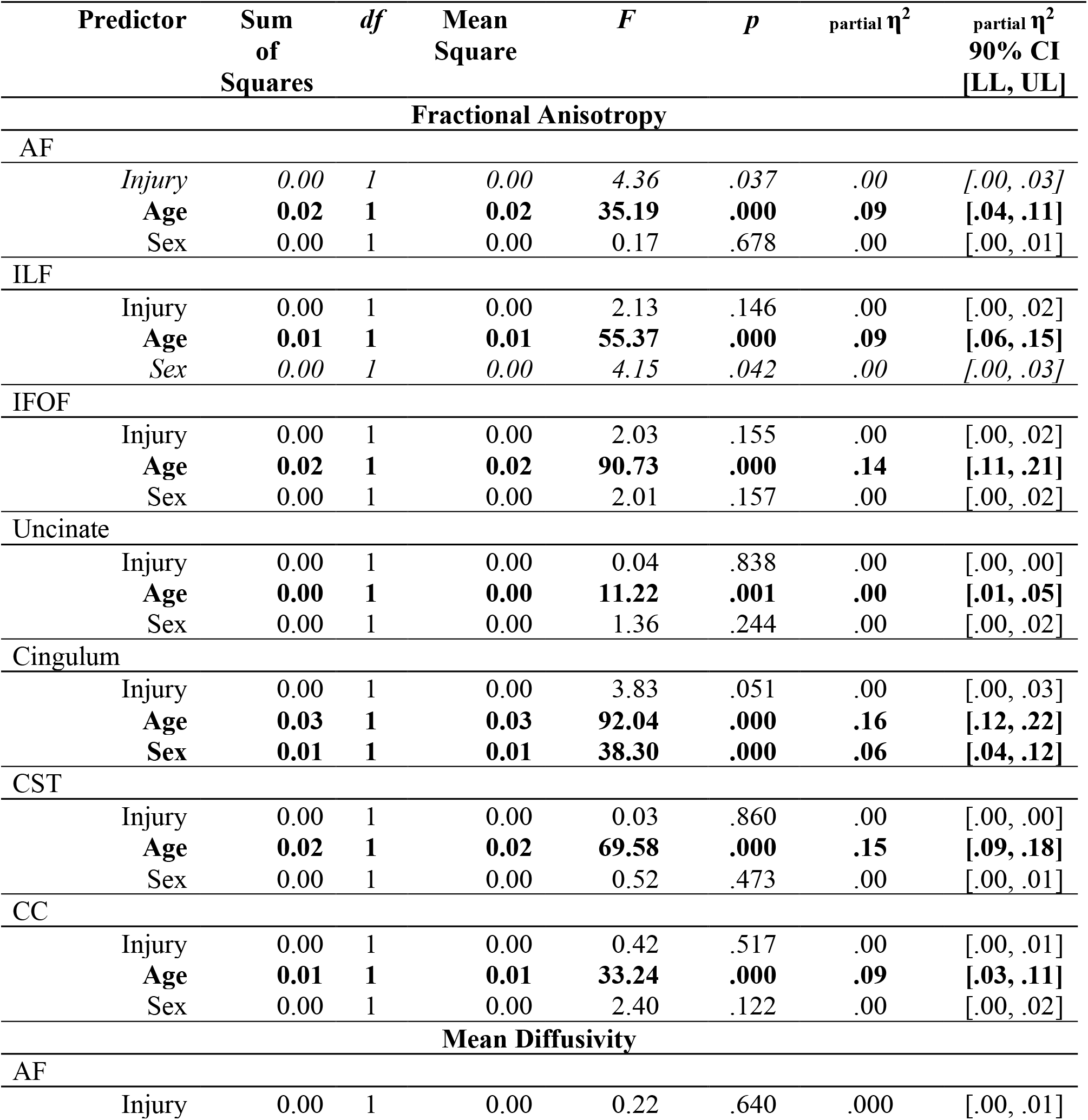
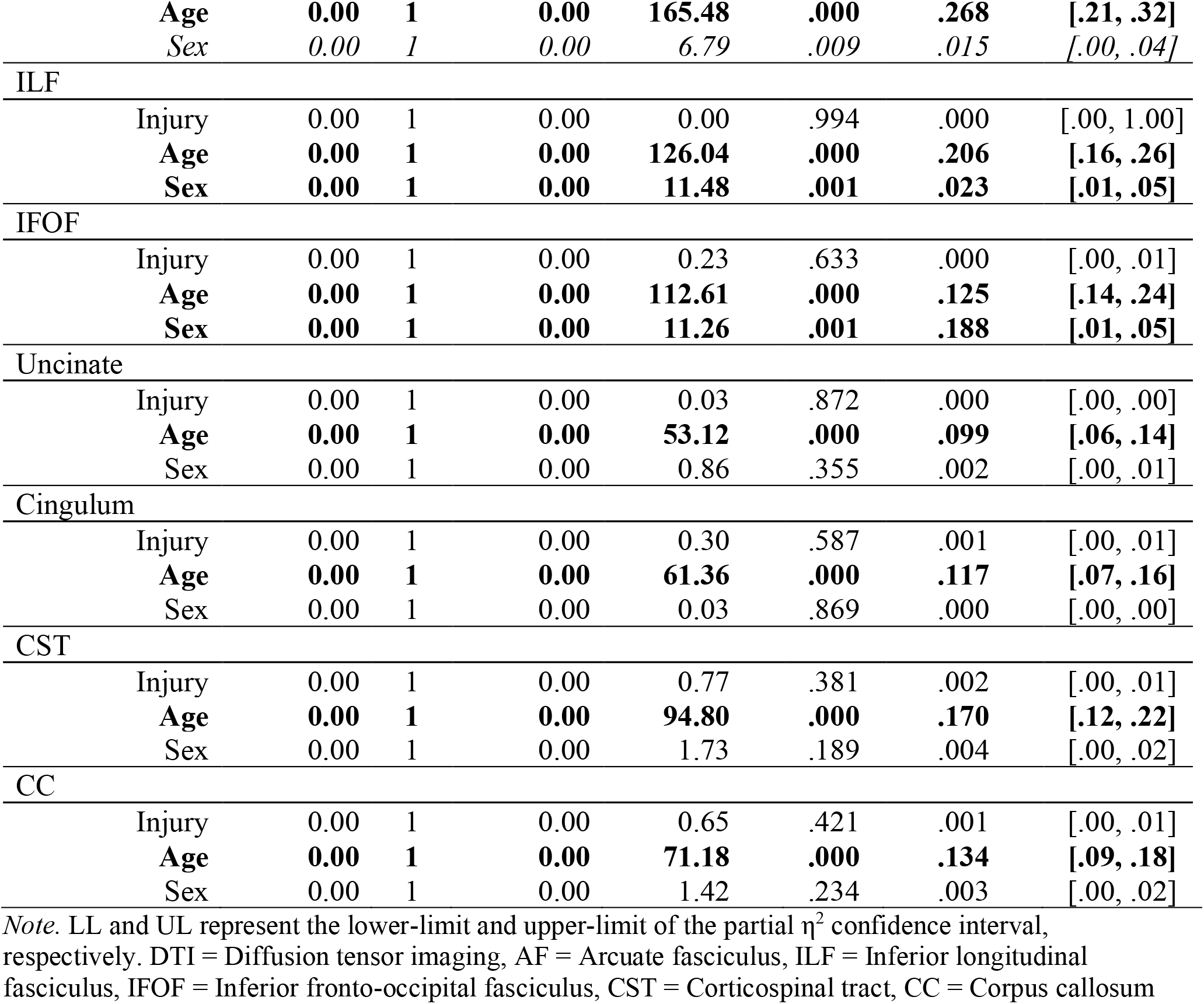
Fixed-Effects ANOVA results for DTI metrics of 7 tracts (left and right combined). Significant p-values after FDR correction are **bolded**, significant p-values that did not survive FDR correction are *italicized*

| Predictor | Sum of Squares | df | Mean Square | F | p | partial $\eta^2$ | partial $\eta^2$ 90% CI [LL, UL] |
| --- | --- | --- | --- | --- | --- | --- | --- |
| **Fractional Anisotropy** |  |  |  |  |  |  |  |
| AF |  |  |  |  |  |  |  |
| *Injury* | *0.00* | *1* | *0.00* | *4.36* | *.037* | *.00* | *[.00, .03]* |
| **Age** | **0.02** | **1** | **0.02** | **35.19** | **.000** | **.09** | **[.04, .11]** |
| *Sex* | *0.00* | *1* | *0.00* | *0.17* | *.678* | *.00* | *[.00, .01]* |
| ILF |  |  |  |  |  |  |  |
| *Injury* | *0.00* | *1* | *0.00* | *2.13* | *.146* | *.00* | *[.00, .02]* |
| **Age** | **0.01** | **1** | **0.01** | **55.37** | **.000** | **.09** | **[.06, .15]** |
| *Sex* | *0.00* | *1* | *0.00* | *4.15* | *.042* | *.00* | *[.00, .03]* |
| IFOF |  |  |  |  |  |  |  |
| *Injury* | *0.00* | *1* | *0.00* | *2.03* | *.155* | *.00* | *[.00, .02]* |
| **Age** | **0.02** | **1** | **0.02** | **90.73** | **.000** | **.14** | **[.11, .21]** |
| *Sex* | *0.00* | *1* | *0.00* | *2.01* | *.157* | *.00* | *[.00, .02]* |
| Uncinate |  |  |  |  |  |  |  |
| *Injury* | *0.00* | *1* | *0.00* | *0.04* | *.838* | *.00* | *[.00, .00]* |
| **Age** | **0.00** | **1** | **0.00** | **11.22** | **.001** | **.00** | **[.01, .05]** |
| *Sex* | *0.00* | *1* | *0.00* | *1.36* | *.244* | *.00* | *[.00, .02]* |
| Cingulum |  |  |  |  |  |  |  |
| *Injury* | *0.00* | *1* | *0.00* | *3.83* | *.051* | *.00* | *[.00, .03]* |
| **Age** | **0.03** | **1** | **0.03** | **92.04** | **.000** | **.16** | **[.12, .22]** |
| **Sex** | **0.01** | **1** | **0.01** | **38.30** | **.000** | **.06** | **[.04, .12]** |
| CST |  |  |  |  |  |  |  |
| *Injury* | *0.00* | *1* | *0.00* | *0.03* | *.860* | *.00* | *[.00, .00]* |
| **Age** | **0.02** | **1** | **0.02** | **69.58** | **.000** | **.15** | **[.09, .18]** |
| *Sex* | *0.00* | *1* | *0.00* | *0.52* | *.473* | *.00* | *[.00, .01]* |
| CC |  |  |  |  |  |  |  |
| *Injury* | *0.00* | *1* | *0.00* | *0.42* | *.517* | *.00* | *[.00, .01]* |
| **Age** | **0.01** | **1** | **0.01** | **33.24** | **.000** | **.09** | **[.03, .11]** |
| *Sex* | *0.00* | *1* | *0.00* | *2.40* | *.122* | *.00* | *[.00, .02]* |
| **Mean Diffusivity** |  |  |  |  |  |  |  |
| AF |  |  |  |  |  |  |  |
| *Injury* | *0.00* | *1* | *0.00* | *0.22* | *.640* | *.000* | *[.00, .01]* |

|  |  |  |  |  |  |  |  |  |
| --- | --- | --- | --- | --- | --- | --- | --- | --- |
|  | **Age** | **0.00** | **1** | **0.00** | **165.48** | **.000** | **.268** | **[.21, .32]** |
|  | *Sex* | *0.00* | *1* | *0.00* | *6.79* | *.009* | *.015* | *[.00, .04]* |
| ILF |  |  |  |  |  |  |  |  |
|  | Injury | 0.00 | 1 | 0.00 | 0.00 | .994 | .000 | [.00, 1.00] |
|  | **Age** | **0.00** | **1** | **0.00** | **126.04** | **.000** | **.206** | **[.16, .26]** |
|  | **Sex** | **0.00** | **1** | **0.00** | **11.48** | **.001** | **.023** | **[.01, .05]** |
| IFOF |  |  |  |  |  |  |  |  |
|  | Injury | 0.00 | 1 | 0.00 | 0.23 | .633 | .000 | [.00, .01] |
|  | **Age** | **0.00** | **1** | **0.00** | **112.61** | **.000** | **.125** | **[.14, .24]** |
|  | **Sex** | **0.00** | **1** | **0.00** | **11.26** | **.001** | **.188** | **[.01, .05]** |
| Uncinate |  |  |  |  |  |  |  |  |
|  | Injury | 0.00 | 1 | 0.00 | 0.03 | .872 | .000 | [.00, .00] |
|  | **Age** | **0.00** | **1** | **0.00** | **53.12** | **.000** | **.099** | **[.06, .14]** |
|  | **Sex** | **0.00** | **1** | **0.00** | **0.86** | **.355** | **.002** | **[.00, .01]** |
| Cingulum |  |  |  |  |  |  |  |  |
|  | Injury | 0.00 | 1 | 0.00 | 0.30 | .587 | .001 | [.00, .01] |
|  | **Age** | **0.00** | **1** | **0.00** | **61.36** | **.000** | **.117** | **[.07, .16]** |
|  | **Sex** | **0.00** | **1** | **0.00** | **0.03** | **.869** | **.000** | **[.00, .00]** |
| CST |  |  |  |  |  |  |  |  |
|  | Injury | 0.00 | 1 | 0.00 | 0.77 | .381 | .002 | [.00, .01] |
|  | **Age** | **0.00** | **1** | **0.00** | **94.80** | **.000** | **.170** | **[.12, .22]** |
|  | **Sex** | **0.00** | **1** | **0.00** | **1.73** | **.189** | **.004** | **[.00, .02]** |
| CC |  |  |  |  |  |  |  |  |
|  | Injury | 0.00 | 1 | 0.00 | 0.65 | .421 | .001 | [.00, .01] |
|  | **Age** | **0.00** | **1** | **0.00** | **71.18** | **.000** | **.134** | **[.09, .18]** |
|  | **Sex** | **0.00** | **1** | **0.00** | **1.42** | **.234** | **.003** | **[.00, .02]** |

*Note.* LL and UL represent the lower-limit and upper-limit of the partial $\eta^2$ confidence interval, respectively. DTI = Diffusion tensor imaging, AF = Arcuate fasciculus, ILF = Inferior longitudinal fasciculus, IFOF = Inferior fronto-occipital fasciculus, CST = Corticospinal tract, CC = Corpus callosum

Before FDR correction, significant group differences between children with mTBI and OI were identified for FA in the bilateral arcuate fasciculus, (*p=*0.037). A significant interaction between injury and age at injury were also identified in FA in the bilateral arcuate fasciculus, (*p =* .023). Post-hoc analysis revealed that lower FA in children with mTBI as compared to OI at younger ages, and higher FA in children with mTBI compared to OI at older ages (*Figure 2a*) in the bilateral AF.

**Figure 2:**
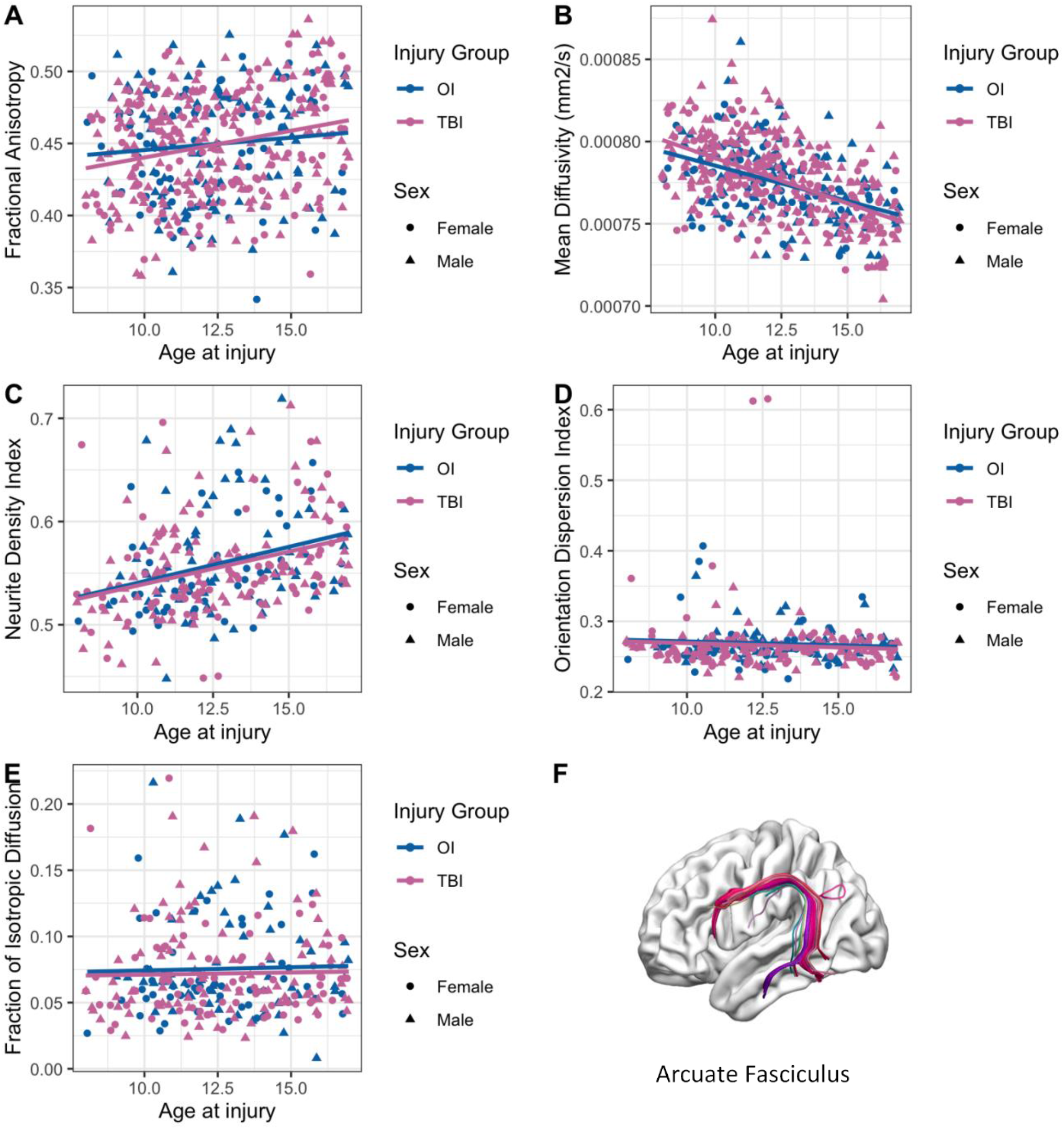
Scatter plots illustrating relation between age and DTI and NODDI metrics of the left/right combined arcuate fasciculus in children with mTBI and OI. Age-related linear correlations were observed for both groups independently: (A) FA, positive; (B) MD, negative; and (C) NDI, positive. No age-related trends were observed in (D) orientation dispersion index or (E) fraction of isotropic diffusion. (F) is an example of the left arcuate fasciculus tract isolated using semi-automated tractography.

In line with previous research^45,48^, FA in all tracts exhibited a significant positive association with age while MD in all tracts had a significant negative association with age (FA increased and MD decreased with age) for both injury groups (*Table 3)*. FA in the cingulum (*p* < .001) and MD in the ILF (*p* = .001), and IFOF (*p* = .001) were significantly associated with sex after FDR correction. (*Table 3)*. Post-hoc analysis revealed higher FA in the cingulum and higher MD in the ILF and IFOF in males as compared to females in the current sample for both injury groups.

### 3.2. NODDI

Results from the univariate ANOVA examining group differences on NODDI metrics (NDI, ODI, FISO) are presented in *Table 4*. No significant differences between children with mTBI and OI were identified on any NODDI metric, before or after FDR correction.

**Table 4.**
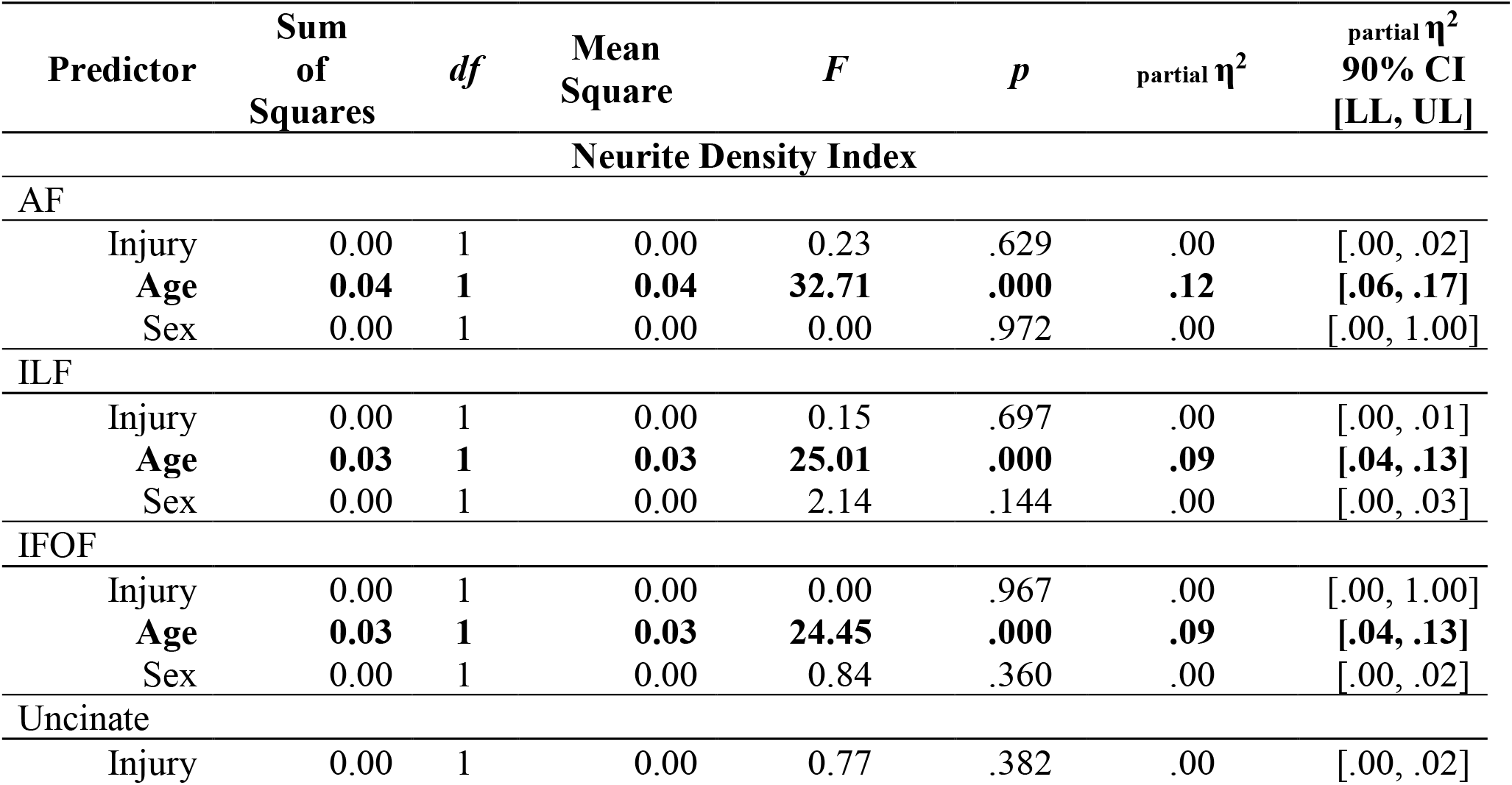
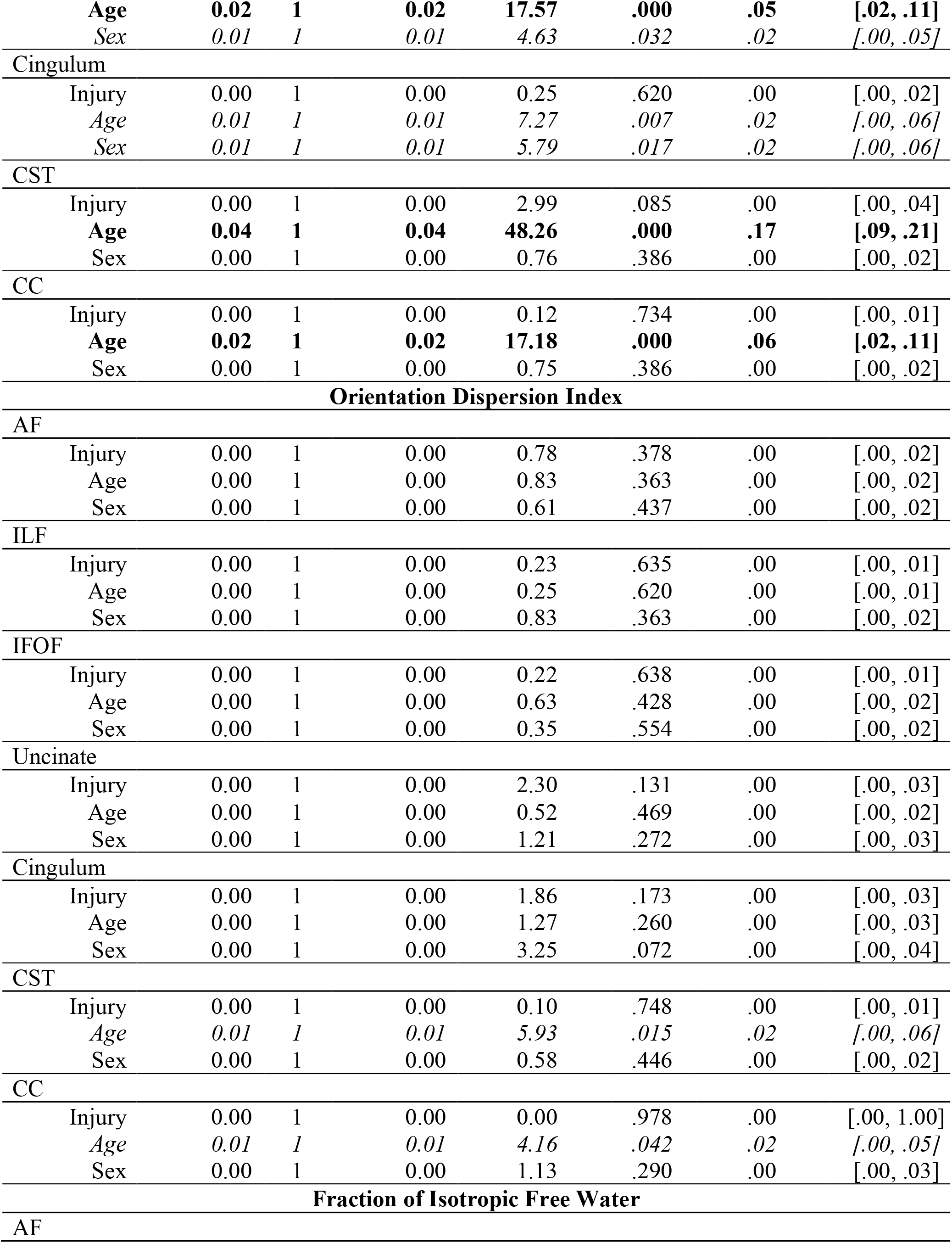
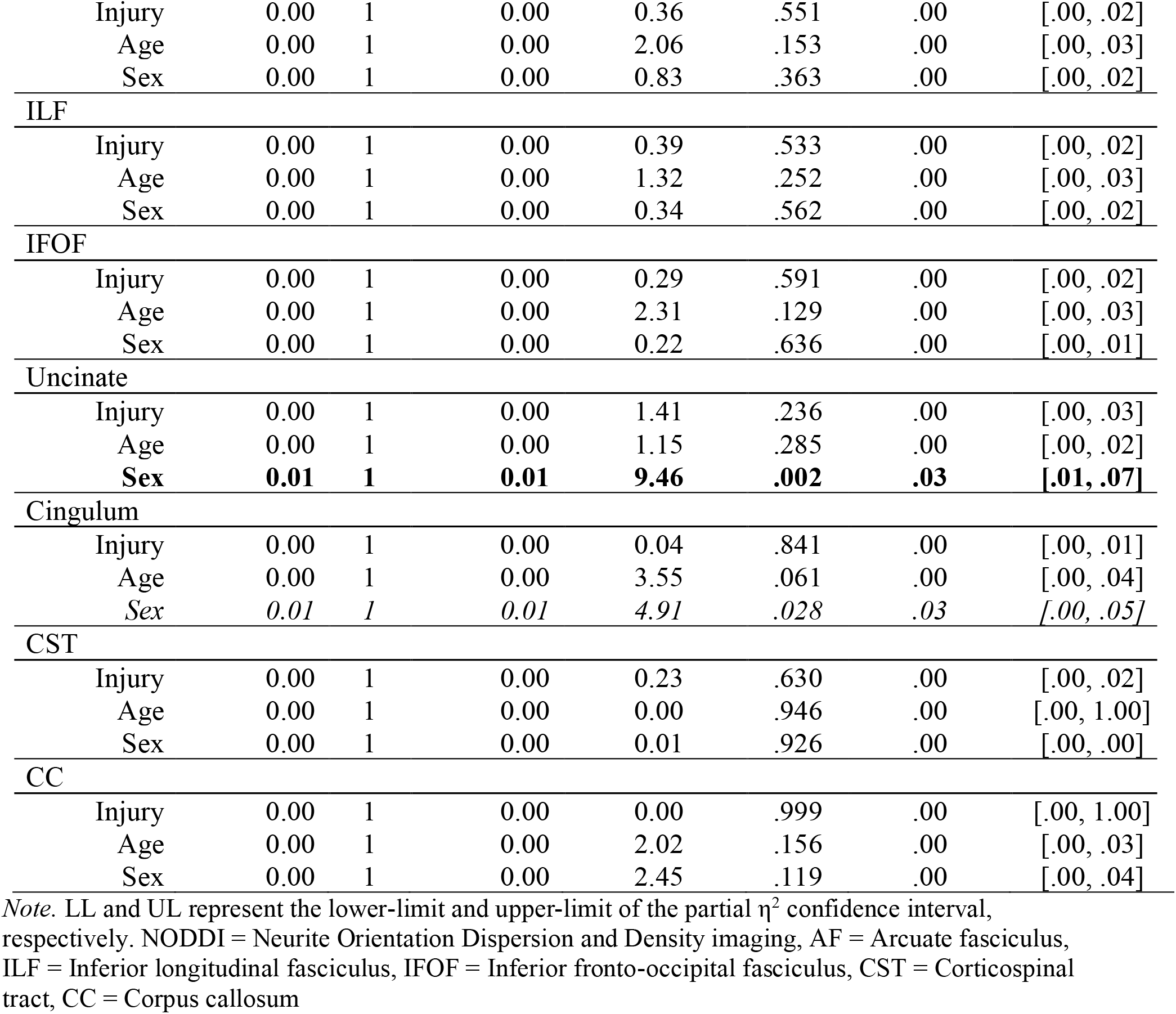
Fixed-Effects ANOVA results for NODDI metrics. Significant p-values after FDR correction are **bolded**, significant p-values that did not survive FDR correction are *italicized*

| Predictor | Sum of Squares | df | Mean Square | F | p | partial $\eta^2$ | partial $\eta^2$ 90% CI [LL, UL] |
| --- | --- | --- | --- | --- | --- | --- | --- |
| Neurite Density Index |  |  |  |  |  |  |  |
| AF |  |  |  |  |  |  |  |
| Injury | 0.00 | 1 | 0.00 | 0.23 | .629 | .00 | [.00, .02] |
| **Age** | **0.04** | **1** | **0.04** | **32.71** | **.000** | **.12** | **[.06, .17]** |
| Sex | 0.00 | 1 | 0.00 | 0.00 | .972 | .00 | [.00, 1.00] |
| ILF |  |  |  |  |  |  |  |
| Injury | 0.00 | 1 | 0.00 | 0.15 | .697 | .00 | [.00, .01] |
| **Age** | **0.03** | **1** | **0.03** | **25.01** | **.000** | **.09** | **[.04, .13]** |
| Sex | 0.00 | 1 | 0.00 | 2.14 | .144 | .00 | [.00, .03] |
| IFOF |  |  |  |  |  |  |  |
| Injury | 0.00 | 1 | 0.00 | 0.00 | .967 | .00 | [.00, 1.00] |
| **Age** | **0.03** | **1** | **0.03** | **24.45** | **.000** | **.09** | **[.04, .13]** |
| Sex | 0.00 | 1 | 0.00 | 0.84 | .360 | .00 | [.00, .02] |
| Uncinate |  |  |  |  |  |  |  |
| Injury | 0.00 | 1 | 0.00 | 0.77 | .382 | .00 | [.00, .02] |

|  |  |  |  |  |  |  |  |  |
| --- | --- | --- | --- | --- | --- | --- | --- | --- |
|  | Age | 0.02 | 1 | 0.02 | 17.57 | .000 | .05 | [.02, .11] |
|  | Sex | 0.01 | 1 | 0.01 | 4.63 | .032 | .02 | [.00, .05] |
| Cingulum |  |  |  |  |  |  |  |  |
|  | Injury | 0.00 | 1 | 0.00 | 0.25 | .620 | .00 | [.00, .02] |
|  | Age | 0.01 | 1 | 0.01 | 7.27 | .007 | .02 | [.00, .06] |
|  | Sex | 0.01 | 1 | 0.01 | 5.79 | .017 | .02 | [.00, .06] |
| CST |  |  |  |  |  |  |  |  |
|  | Injury | 0.00 | 1 | 0.00 | 2.99 | .085 | .00 | [.00, .04] |
|  | Age | 0.04 | 1 | 0.04 | 48.26 | .000 | .17 | [.09, .21] |
|  | Sex | 0.00 | 1 | 0.00 | 0.76 | .386 | .00 | [.00, .02] |
| CC |  |  |  |  |  |  |  |  |
|  | Injury | 0.00 | 1 | 0.00 | 0.12 | .734 | .00 | [.00, .01] |
|  | Age | 0.02 | 1 | 0.02 | 17.18 | .000 | .06 | [.02, .11] |
|  | Sex | 0.00 | 1 | 0.00 | 0.75 | .386 | .00 | [.00, .02] |
| Orientation Dispersion Index |  |  |  |  |  |  |  |  |
| AF |  |  |  |  |  |  |  |  |
|  | Injury | 0.00 | 1 | 0.00 | 0.78 | .378 | .00 | [.00, .02] |
|  | Age | 0.00 | 1 | 0.00 | 0.83 | .363 | .00 | [.00, .02] |
|  | Sex | 0.00 | 1 | 0.00 | 0.61 | .437 | .00 | [.00, .02] |
| ILF |  |  |  |  |  |  |  |  |
|  | Injury | 0.00 | 1 | 0.00 | 0.23 | .635 | .00 | [.00, .01] |
|  | Age | 0.00 | 1 | 0.00 | 0.25 | .620 | .00 | [.00, .01] |
|  | Sex | 0.00 | 1 | 0.00 | 0.83 | .363 | .00 | [.00, .02] |
| IFOF |  |  |  |  |  |  |  |  |
|  | Injury | 0.00 | 1 | 0.00 | 0.22 | .638 | .00 | [.00, .01] |
|  | Age | 0.00 | 1 | 0.00 | 0.63 | .428 | .00 | [.00, .02] |
|  | Sex | 0.00 | 1 | 0.00 | 0.35 | .554 | .00 | [.00, .02] |
| Uncinate |  |  |  |  |  |  |  |  |
|  | Injury | 0.00 | 1 | 0.00 | 2.30 | .131 | .00 | [.00, .03] |
|  | Age | 0.00 | 1 | 0.00 | 0.52 | .469 | .00 | [.00, .02] |
|  | Sex | 0.00 | 1 | 0.00 | 1.21 | .272 | .00 | [.00, .03] |
| Cingulum |  |  |  |  |  |  |  |  |
|  | Injury | 0.00 | 1 | 0.00 | 1.86 | .173 | .00 | [.00, .03] |
|  | Age | 0.00 | 1 | 0.00 | 1.27 | .260 | .00 | [.00, .03] |
|  | Sex | 0.00 | 1 | 0.00 | 3.25 | .072 | .00 | [.00, .04] |
| CST |  |  |  |  |  |  |  |  |
|  | Injury | 0.00 | 1 | 0.00 | 0.10 | .748 | .00 | [.00, .01] |
|  | Age | 0.01 | 1 | 0.01 | 5.93 | .015 | .02 | [.00, .06] |
|  | Sex | 0.00 | 1 | 0.00 | 0.58 | .446 | .00 | [.00, .02] |
| CC |  |  |  |  |  |  |  |  |
|  | Injury | 0.00 | 1 | 0.00 | 0.00 | .978 | .00 | [.00, 1.00] |
|  | Age | 0.01 | 1 | 0.01 | 4.16 | .042 | .02 | [.00, .05] |
|  | Sex | 0.00 | 1 | 0.00 | 1.13 | .290 | .00 | [.00, .03] |
| Fraction of Isotropic Free Water |  |  |  |  |  |  |  |  |
| AF |  |  |  |  |  |  |  |  |
| Injury | 0.00 | 1 | 0.00 | 0.36 | .551 | .00 | [.00, .02] |
| Age | 0.00 | 1 | 0.00 | 2.06 | .153 | .00 | [.00, .03] |
| Sex | 0.00 | 1 | 0.00 | 0.83 | .363 | .00 | [.00, .02] |
| ILF |  |  |  |  |  |  |  |
| Injury | 0.00 | 1 | 0.00 | 0.39 | .533 | .00 | [.00, .02] |
| Age | 0.00 | 1 | 0.00 | 1.32 | .252 | .00 | [.00, .03] |
| Sex | 0.00 | 1 | 0.00 | 0.34 | .562 | .00 | [.00, .02] |
| IFOF |  |  |  |  |  |  |  |
| Injury | 0.00 | 1 | 0.00 | 0.29 | .591 | .00 | [.00, .02] |
| Age | 0.00 | 1 | 0.00 | 2.31 | .129 | .00 | [.00, .03] |
| Sex | 0.00 | 1 | 0.00 | 0.22 | .636 | .00 | [.00, .01] |
| Uncinate |  |  |  |  |  |  |  |
| Injury | 0.00 | 1 | 0.00 | 1.41 | .236 | .00 | [.00, .03] |
| Age | 0.00 | 1 | 0.00 | 1.15 | .285 | .00 | [.00, .02] |
| Sex | **0.01** | **1** | **0.01** | **9.46** | **.002** | **.03** | **[.01, .07]** |
| Cingulum |  |  |  |  |  |  |  |
| Injury | 0.00 | 1 | 0.00 | 0.04 | .841 | .00 | [.00, .01] |
| Age | 0.00 | 1 | 0.00 | 3.55 | .061 | .00 | [.00, .04] |
| Sex | *0.01* | *1* | *0.01* | *4.91* | *.028* | *.03* | *[.00, .05]* |
| CST |  |  |  |  |  |  |  |
| Injury | 0.00 | 1 | 0.00 | 0.23 | .630 | .00 | [.00, .02] |
| Age | 0.00 | 1 | 0.00 | 0.00 | .946 | .00 | [.00, 1.00] |
| Sex | 0.00 | 1 | 0.00 | 0.01 | .926 | .00 | [.00, .00] |
| CC |  |  |  |  |  |  |  |
| Injury | 0.00 | 1 | 0.00 | 0.00 | .999 | .00 | [.00, 1.00] |
| Age | 0.00 | 1 | 0.00 | 2.02 | .156 | .00 | [.00, .03] |
| Sex | 0.00 | 1 | 0.00 | 2.45 | .119 | .00 | [.00, .04] |

*Note.* LL and UL represent the lower-limit and upper-limit of the partial $\eta^2$ confidence interval, respectively. NODDI = Neurite Orientation Dispersion and Density imaging, AF = Arcuate fasciculus, ILF = Inferior longitudinal fasciculus, IFOF = Inferior fronto-occipital fasciculus, CST = Corticospinal tract, CC = Corpus callosum

In line with previous research^49–51^, NDI in all tracts was significantly positively associated with age in all tracts examined, whereas no significant associations were identified between ODI and age (*Table 4)*. NDI in the uncinate fasciculus (*p* = .002) was significantly associated with sex, these results survived FDR correction (*Table 3)*. Post-hoc analysis revealed higher NDI in males in the uncinate fasciculus as compared to females in the current sample.

## 4. Discussion

In our large prospective study of children with mTBI, we found no significant differences in diffusion parameters within white matter tracts at the post-acute stage of injury compared to children with orthopedic injuries. This suggests that changes to brain microstructure may not be apparent in the first few weeks following mTBI.

Some previous studies of pediatric mild TBI have identified widespread white matter alterations at the post-acute period of injury,^19,52,53^ whereas others have reported no significant differences in DTI metrics between children with mild TBI and controls.^54,55^ This heterogeneity in findings can likely be attributed to small sample sizes (n = 6 - 83),^17,31,53^ limited (14 - 17 years)^20^ or very large (10 – 38 years) age ranges,^56,19,52,55^ different comparison groups between studies (OI versus uninjured controls),^19,30,31,52^ and differing image analysis techniques (voxel-wise,^16,57^ tract based special statistics,^56^ whole brain histogram analysis,^58^ deterministic tractography,^59,60^ probabilistic tractography^61^). To address the above-mentioned factors that lead to non-uniformity in studies of pediatric mTBI, the current study included a sample size large enough to provide sufficient power to detect small effects (*f* = .10), an age range (8-16.99 years) old enough to ensure reliable self-reporting of symptoms and successful completion of MRI scans while limiting developmental heterogeneity, and an OI comparison group to control for premorbid behavioral differences between children with mTBI and uninjured controls as well as post-injury factors such as stress, pain, and medication effects.^55^ Therefore, our findings suggest that the lack of findings here in multiple brain areas may represent a true lack of measurable changes in white matter structure in children with mTBI at the post-acute injury stage.

NODDI metrics are more strongly associated with age-related changes in white matter in both injury groups, compared to the more standard DTI metrics of FA and MD^50,62^. In line with this, we identified a positive association of NDI with age in all tracts examined here. However, no differences in NODDI metrics were identified between children with mild TBI versus OI in any of the tracts examined here. Previous NODDI studies of mild TBI in adults have reported lower NDI and higher FISO after mTBI as compared to OI 2-weeks post injury, with decreases in NDI longitudinally.^30,31,63^ MTBI leads to a neurometabolic cascade beginning with ionic flux and glutamate release at early stages of injury, followed by cytoskeletal damage, axonal dysfunction, and altered neurotransmission, which may lead to inflammation and possible cell death.^64^ The trajectory of these metabolic changes in children differs from that in adults due to higher vulnerability of the younger brain to biomechanical effects after concussion,^65^ making the trajectory of white matter changes post-injury different between the pediatric and adult population, which may explain the lack of differences between the two groups here. Alternatively, in children, white matter alterations after mTBI may not be evident post-acutely and rather may evolve over the course of injury and be observed at a later stage in injury, indicating a need for longitudinal follow-up studies.

An important consideration while interpreting the lack of differences in white matter microstructure between the injury groups in this study is the severity or presence of PCS within in the mTBI group. There may be a subgroup within the mTBI group with more severe PCS as compared to the rest of the mTBI group, which may exhibit altered white matter microstructure at the post-acute stage of injury, but these differences may be lost due to the inclusion of the whole mTBI sample, instead of a subset of children. Therefore, examining PCS and their association with white matter microstructure in future studies is an important future direction.

Pediatric mild TBIs have multiple mechanisms of injury^66^ and lead to diffuse axonal injuries, causing widespread disruptions in brain white matter connectivity.^67,68^ A graph theory or connectome-based approach to map white matter networks after injury is well suited to characterize these disruptions. Therefore, future studies employing network-based approaches in addition to the currently used structural metrics may yield further insight into pediatric mTBI.

### 4.1. Limitations

The results from this study should be viewed in light of some limitations. The data were collected at multiple sites across Canada to enable the recruitment of a large study sample. Protocols were standardized, but differences in scan parameters, scanner manufacturers, and sites may introduce confounding factors to images.^69^ To account for this, the current study included the scanner at which the image was acquired as a covariate. We did not obtain pre-injury baseline scans, this is due to methodological difficulties; therefore, it cannot be concluded whether the DTI and NODDI metrics observed in this study were a result of injury as compared to pre-injury factors. However, using children with mild OI as the comparison group enabled us to to control for factors that predispose children to injuries, as well as biological and behavioral characteristics (brain changes, pain, etc.) that are caused by injuries in general.

### 4.2. Conclusions

The current study extends our understanding of white matter microstructure at the post-acute stage of pediatric mild TBI. White matter characteristics post-acutely after injury were similar between children with mTBI and OI, suggesting that differences in the biophysical properties of white matter tracts are not apparent at the post-acute stage of injury. Using an OI comparison group in the current study enabled us to control for some common confounding factors that blur comparisons of mTBI to healthy children. White matter differences following mTBI in children may emerge over time, so longitudinal studies are needed, as are analyses of the association between neuroimaging data and PCS.

## Data Availability

The data will be available upon reasonable request

